# Assessing Risk Factors for Cognitive Decline Using Electronic Health Records: A Scoping Review

**DOI:** 10.1101/2023.11.06.23298163

**Authors:** Liqin Wang, Richard Yang, Ziqin Sha, Anna Maria Kuraszkiewicz, Conrad Leonik, Li Zhou, Gad A. Marshall

## Abstract

**Background:** Electronic health records (EHR) provide rich, diverse, longitudinal real-world data, offering valuable opportunities to study antecedent risk factors for cognitive decline. However, the extent to which such records have been utilized to understand the risk factors of cognitive decline remains unclear.

**Methods:** We conducted a systematic review following PRISMA guidelines. We retrieved relevant articles published between January 1, 2010, and April 30, 2023, from PubMed, Web of Science, and CINAHL. Inclusion criteria were studies utilizing EHR to study risk factors for cognitive decline, including subjective cognitive decline, mild cognitive impairment, and dementia. Each article was screened by at least two reviewers. Data elements were manually extracted based on a predefined schema. The studied risk factors were reported.

**Results:** Of 1,593 articles identified, 80 research articles were selected. Seventy (87.5%) were retrospective cohort studies, 53 (66.3%) used datasets with greater than 10,000 patients, and 69 (86.3%) used EHR datasets from the United States or United Kingdom. Furthermore, 39 (48.8%) targeted medical conditions, 23 (28.8%) related to medical interventions, and 14 (17.5%) related to lifestyle, socioeconomic status, and environmental factors. The majority of studied medical conditions were associated with an elevated risk of cognitive decline, whereas medical interventions addressing these conditions often reduced the risk.

**Conclusion:** EHRs have significantly advanced our understanding of medical conditions, interventions, lifestyle, socioeconomic status, and environmental factors related to the risk of cognitive decline.

**Highlights:**

1. Electronic health records are increasingly analyzed to discern dementia risk factors.
2. Eighty articles have been published and most of them were retrospective cohort studies.
3. Most risk factors addressed were related to medical conditions and interventions.
4. A few articles studied lifestyle, socioeconomic status, and environmental factors.

## INTRODUCTION

Alzheimer’s disease (AD), characterized by chronic or progressive cognitive and functional decline, is a major cause of disability and dependency among older adults, causing great challenges for public health worldwide.^1^ Based on the degree of cognitive impairment, AD is often divided into three stages: the preclinical stage, characterized by normal cognitive ability with or without subtle cognitive concerns but with biological evidence of underlying AD, prodromal stage, characterized by mild cognitive impairment (MCI), and dementia stage with significant functional impairment interfering with daily activities.^2, 3^ In 2023, it is estimated that 6.7 million Americans are living with AD at the stage of dementia in the US, and this number is expected to increase to 88 million by the year of 2050.^4^ This alarming statistic is not only significant in financial terms, but also underscores the profound impact on the affected individuals and families and the healthcare system at large. Understanding the risk factors that might contribute to dementia and identifying potential prevention or treatment strategies that might reduce the risk are of utmost importance.

While traditional research methodologies, focusing on in-depth studies of small cohorts over extended periods^5, 6^ have indeed provided insights into certain aspects of the disease, there is a growing consensus in the scientific community about the need to broaden our scope and explore larger, more diverse datasets.^2^ The widespread adoption of electronic health records (EHRs) over recent decades has yielded a vast amount of longitudinal patient data. By sifting through these real-world datasets, we can gain deeper insights into the onset and evolution of AD and related dementias (ADRD), especially among populations that have been consistently engaged with the healthcare system. EHRs can be valuable in identifying potential risk factors for ADRD that might be missed in smaller convenience sample datasets. Moreover, they can highlight interventions that target certain medical problems that potentially affect the risk of dementia, particularly during early stages such as preclinical AD and MCI. However, the extent to which EHR data have been harnessed for such research remains unclear.

Previous literature review articles have primarily focused on specific areas related to ADRD risk.^7-9^ However, to our knowledge, none have specifically addressed how studies are using EHR data for analyzing ADRD risks. The current study aims to highlight the risk factors being studied through large-scale EHR data. Specifically, with this systematic review, we aimed to comprehensively consolidate existing literature on the use of EHR data in studying risk factors for cognitive decline ranging from MCI to dementia, and to identify unexplored areas of research for potential future investigations.

## METHODS

### Search Strategy

This systematic review followed the Preferred Reporting Items for Systematic Reviews and Meta-Analysis (PRISMA) guidelines.^10^ We employed Boolean search strategies to identify studies written in English and published between January 1, 2010 and April 30, 2023. The databases used were PubMed, Web of Science, and CINAHL. The keyword-based search of the databases consisted of two parts, one for the outcomes and the other for the EHR. For the outcome, we aimed to encompass studies targeting all stages of cognitive impairment; therefore, we did not only include dementia as the keyword, but also relevant words related to the early stages of cognitive decline, such as MCI and normal cognition. The final included keywords were “dementia”, “mild cognitive impairment”, “cognitive dysfunction”, “preclinical Alzheimer”, “cognitive decline”, “normal cognition”, and “Alzheimer”. For the EHR, we included keywords like “electronic health records”, “electronic medical records” and “EHR”. The specific queries for individual databases can be found in **eTable 1**.

### Study Selection

We restricted our selection to studies that utilized EHR data as an essential data source to explore the association of potential risk factors with dementia outcomes. Studies were excluded if they were review articles without original data, were written in a non-English language, focused on patients who already had cognitive impairment at the time of enrollment, had an extremely small sample size (n<100) or short follow-up time for outcomes (<1 year), were non-epidemiology study (e.g., focused on algorithm evaluation), or were of low quality with missing or unclear components (e.g., unclear diagnostic criteria for outcomes).

### Screening Process

Abstracts from the search results were retrieved after removing duplicates and those deemed ineligible by automation tools. Two reviewers independently assessed the titles and abstracts of the remaining articles against the inclusion and exclusion criteria. Discrepancies were resolved through discussion until consensus was reached. Articles that passed the title and abstract screening then underwent full-text screening. Two reviewers independently analyzed these articles, with a senior reviewer addressing any disagreements.

### Data Extraction

From these articles, we extracted articles assessing risk factors for the onset of cognitive decline, including MCI, AD, and other dementias. The methodological quality was assessed, and a data extraction schema was developed, based on the STROBE checklist for observational studies.^11^ Extracted data elements included article information (authors and year), objectives, study design (e.g., cohort or case-control), sample size, age of the study participants, follow-up duration, data sources, explored risk factors, confounding variables, outcomes and measurement, statistical methods, main findings, and interpretation. Each article was assigned to two reviewers who independently extracted the information. Discrepancies were addressed through discussions or consultation with a third reviewer.

### Article Classification

After we extracted specific risk factors from individual articles, we grouped them into major categories, including medical conditions, medical interventions, lifestyle, socioeconomic, psychosocial, and environmental factors. These major categories were further divided into subcategories, such as cardiovascular and metabolic conditions and psychiatric conditions under medical conditions. Since some articles investigated multiple risk factors, a single article might fall under more than one major category or subcategory.

## RESULTS

**Figure 1** shows the PRISMA flow diagram. The initial search resulted in 1,593 articles, comprising 565 from PubMed, 538 from Web of Science, and 490 from CINAHL. A total of 496 duplicate articles were removed. Automated tools marked 74 articles as ineligible; these consisted of 3 case reports, 28 review articles, and 43 articles lacking abstracts. An additional 95 records were eliminated, including 42 datasets, 25 preprints, 13 articles without authors, 9 patents, 3 generic publications, 2 books, and 1 thesis. Subsequently, 832 articles were excluded during the title and abstract screening phase for not meeting the inclusion and exclusion criteria. The remaining 96 articles underwent full-text screening by at least two reviewers. After excluding additional 16 articles for various reasons–such as not using EHR as a major data source, being an atypical epidemiology study, or having extremely small sample size–80 articles remained and were included in the final analysis. A detailed list of these articles and extracted data elements are available in **eTable 2** in the supplement.

**Figure 1.**
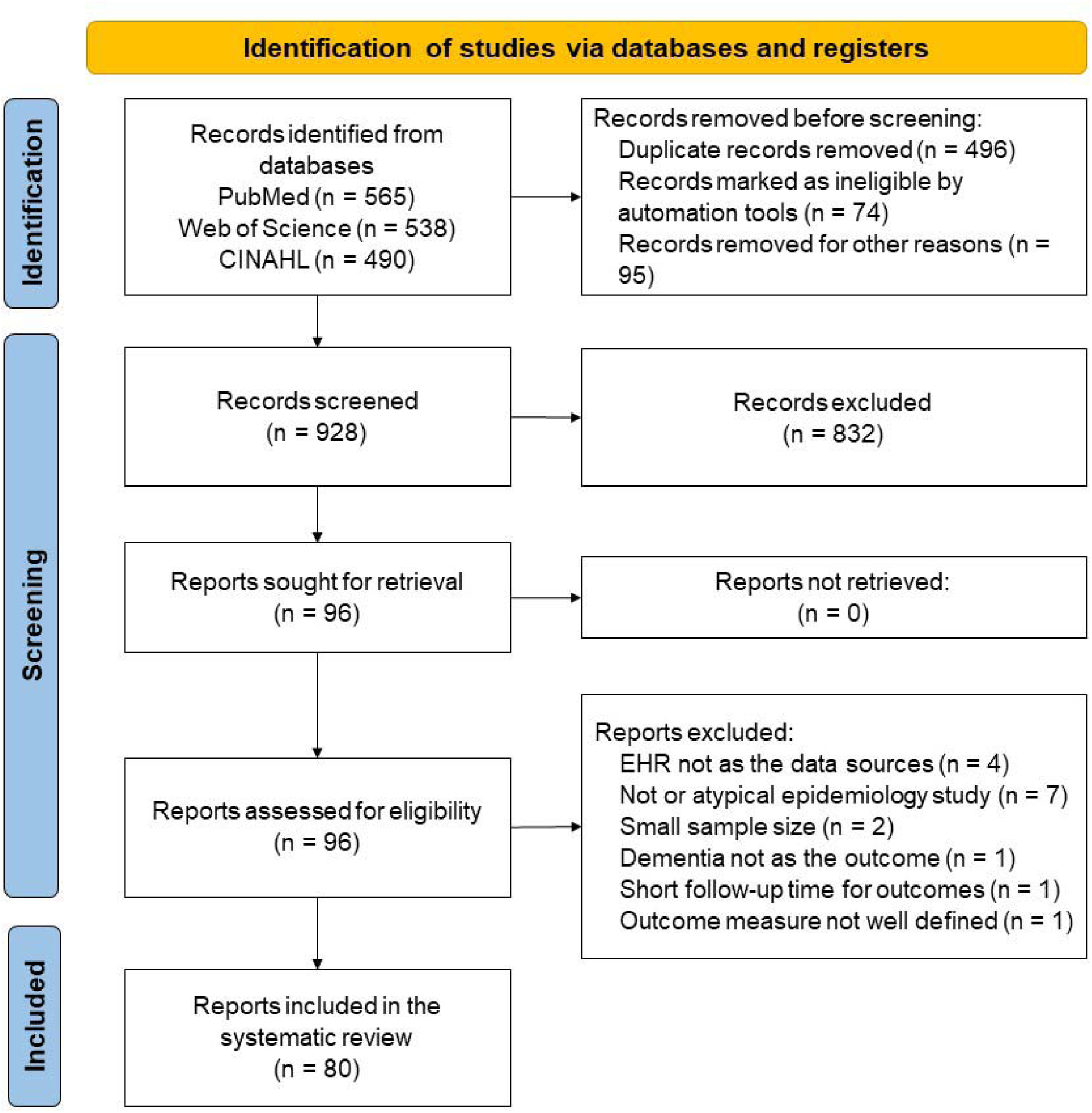
PRISMA flow diagram

### Research Trend Over Time

**Figure 2** illustrates the spread of the articles analyzed based on their year of publication. The chart reveals an increasing number of publications related to our topic over the past decade, indicating a rising trend in the use of EHR data to examine ADRD risk factors. While our search criteria included articles published between 2010 and 2023, all the final included articles were published after 2014. Over one-quarter (n=22, 27.5%) of the articles were published in the year 2022. Our search was conducted up to April 2023; therefore, the total number of articles for that year does not represent the full annual count.

**Figure 2.**
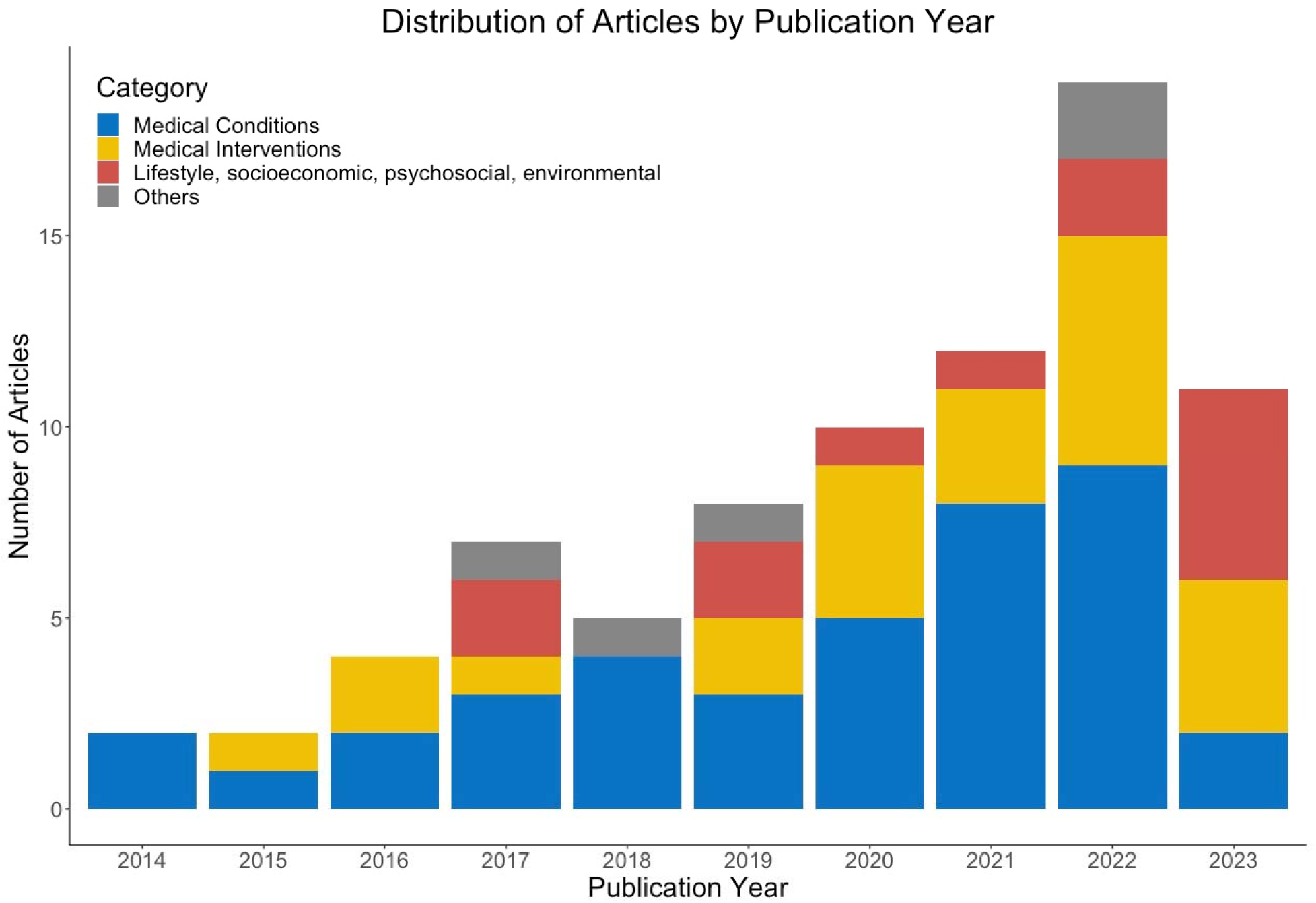
Distribution of articles by publication years, by type of risk factors

### Study Design

Of the 80 articles reviewed, 77(96.3%) were longitudinal studies: 70 were cohort studies, six were case-control studies, and one was a randomized control trial. The remaining three were cross-sectional studies. The longitudinal studies had a median EHR duration of 16 years. The EHR duration for each study was determined from the initial year to the final year of the EHR records utilized, regardless of the actual follow-up time for individual patients. Among them, 16 studies (20%) had a EHR duration of under 10 years, 39 studies (48.8%) ranged between 10 and 20 years, and 22 studies (27.5%) had data duration exceeding 20 years.

### Methods for Statistical Analyses

In the statistical analysis, 76.3% of the studies (n=61) predominantly used survival analysis to model and estimate the outcome-free times and identify various risk or protective factors. Among these, a substantial majority (n=54, 88.5%) chose the Cox proportional-hazards regression model,^12^ while a minority (n=13, 21%) used the Fine-Gray model,^13^ with some studies employing both. The Fine-Gray model was selected specifically for its capability to handle competing risks like death. Additional statistical analysis methods employed included logistic regression, chi-squared test, and analysis of variance (ANOVA).

### EHR Datasets Sources

The included articles utilized various EHR datasets from different geographical locations to examine risk factors for ADRD. A majority of these studies used EHR data from the United States, constituting 46.3% (n=37) of the total. The most commonly used EHR database is that of Kaiser Permanente, with 11 studies, followed by the Veterans Health Administration (VHA), with 6 studies. The remaining 21 articles used databases from other large healthcare systems across the US, as well as some commercial databases, such as TriNetX,^14-16^ IBM Explorys,^17^ and Optum.^18^ Datasets from the United Kingdom (UK) made up 40% (n=32) of the studies. Cohorts from the Whitehall II study^19-24^ (n=8) and UK biobank^25-31^ (n=7) were frequently analyzed for dementia risk factors, during which the datasets were linked to various EHR datasets across the UK, including the Hospital Episode Statistics in England,^32^ the Scottish Morbidity Record data in Scotland,^33^ and the Patient Episode Database in Wales,^25, 26, 31^ to extract a more detailed medical history of patients. Other commonly used databases in the UK studies included the Clinical Practice Research Datalink (n=6)^34-38^ and the Health Improvement Network (THIN) (n=4).^39-41^ Additional countries of databases represented in this review included Australia (n=3),^42-44^ China (n=3),^45-47^ Denmark (n=3),^41, 48, 49^ the Netherlands (n=3),^41, 49, 50^ Taiwan (n=2),^51, 52^ Canada (n=2),^53, 54^ and Sweden (n=2).^55, 56^

### EHR Dataset Sample Size

The studies utilized datasets with sample sizes ranging from several hundred to millions of patients. Only one study used a sample of less than 1000 patients.^53^ Twenty-six (32.5%) studies used datasets ranging from one thousand up to 10 thousand patients; 46 (57.5%) studies had datasets between 10 thousand and one million; and 7 (8.8%) studies used datasets with more than one million patients.

### Outcomes and Measurements

Most studies (n=67) examined multiple dementia subtypes, including AD, vascular dementia, Lewy body dementia (LBD), frontotemporal dementia (FTD), and mixed dementia. While the specific dementia subtypes considered in these studies varied, all studies included AD. Additionally, nine studies exclusively analyzed AD. To define those outcomes, the majority of these studies used codes from standard coding systems, including ICD codes, Read codes, SNOMED-CT, or a combination thereof. ICD codes were the most prevalent, featuring in 81.3% (n=65) of the studies, followed by Read codes in 11.3% (n=9). In addition to these coding systems, several other methods were used to define the outcomes, including prescriptions for dementia medications,^18, 45, 47, 54^ cognitive function tests,^27, 53, 57^ referencing the Diagnostic and Statistical Manual of Mental Disorders (Fourth Edition),^50, 58, 59^ conducting screening interviews,^43, 57^ and neuroimaging.^27^

### Risk and Protective Factors

We summarized the risk factors analyzed in the reviewed articles. For medical conditions and interventions, we have further classified them into broad disease categories (**Table 1**). For the remaining risk factors, we classified them under the categories of lifestyle, socioeconomic, environmental, and others (**Table 2**).

**Table 1.**
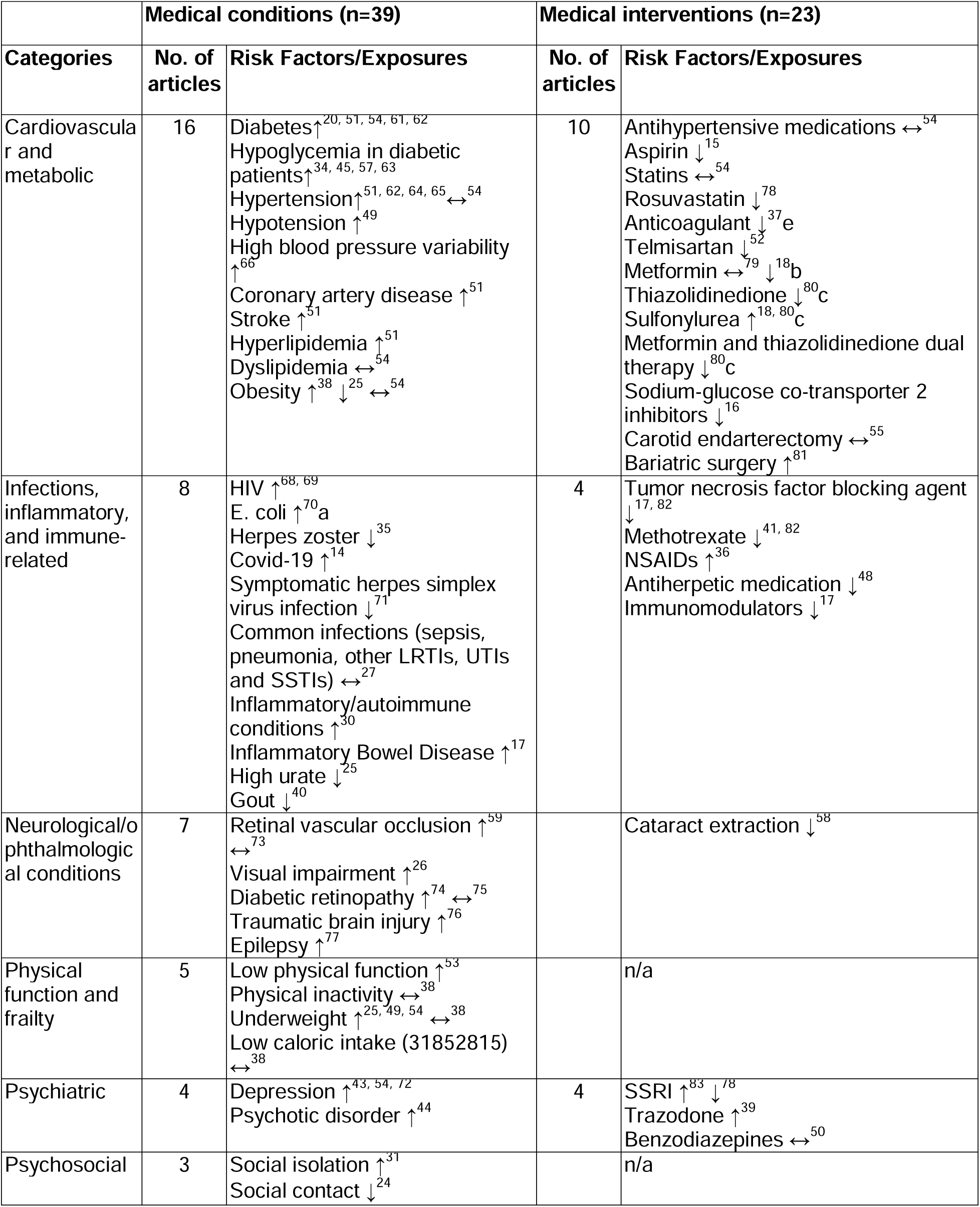
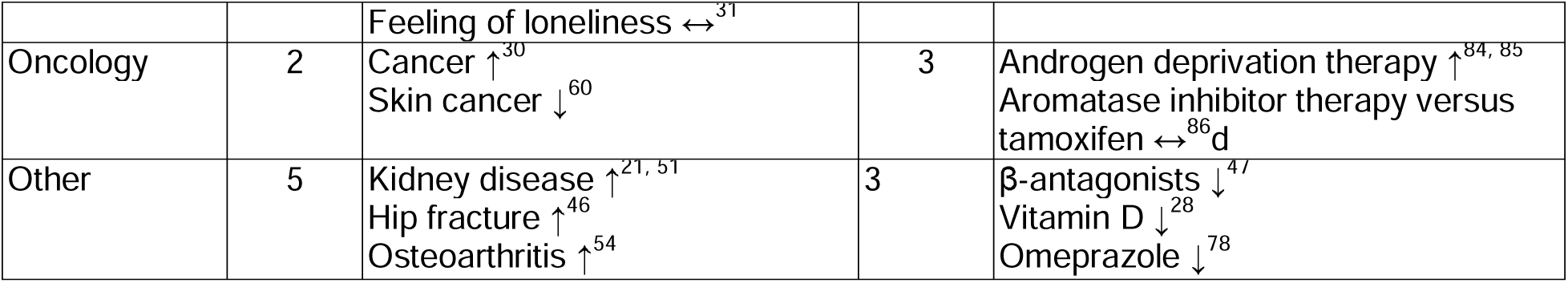
Summary of medical conditions and interventions from EHR-based studies in related to the risk of Alzheimer’s disease and related dementias.

**Table 2.** Summary of lifestyles, socioeconomic, psychosocial, environmental and other factors from EHR-based studies in related to the risk of Alzheimer’s disease and related dementias.

| Categories | No. of articles | Risk Factors/Exposures |
| --- | --- | --- |
| **Lifestyle, socioeconomic, psychosocial, environmental risk factors (n=14)** |  |  |
| Lifestyle | 5 | Diet ↔<sup>22, 87</sup><br>Healthy lifestyle ↓<sup>29</sup><br>Smoking ↑<sup>54</sup><br>Alcohol consumption ↑<sup>56</sup> |
| Socioeconomic | 5 | High education ↓<sup>88, 89</sup> ↔<sup>23</sup><br>Neighborhood disadvantage ↑<sup>54, 90</sup><br>Low occupational position ↑<sup>23</sup> |
| Psychosocial | 3 | Social isolation ↑<sup>31</sup><br>Social contact ↓<sup>24</sup><br>Feeling of loneliness ↔<sup>31</sup> |
| Environmental | 3 | Birth in high stroke mortality states ↑<sup>91</sup><br>Agent orange ↑<sup>92</sup><br>Lithium level in drinking water ↑<sup>93</sup> |
| **Others (n=7)** |  |  |
| Others | 6 | Plasma protein ↑<sup>19</sup><br>Lower testosterone ↑<sup>42</sup><br>Higher brain age ↑<sup>106</sup><br>Low Childhood IQ ↑<sup>107</sup><br>ICU admission ↑<sup>108</sup><br>Hispanic race ↑<sup>62</sup><br>Sex ↔<sup>54</sup><br>CRP genotype ↔<sup>30</sup><br>Apolipoprotein E (APOE) genotype ↔<sup>30</sup> |

**Abbreviations:** HIV, human immunodeficiency virus; LRTIs, lower respiratory tract infections; UTIs, urinary tract infections SSTIs, soft tissue infections; BMI, body mass index; ICU, intensive care unit; CRP, c-reactive protein; APOE, apolipoprotein E.
↑ indicates increased risk for cognitive decline
↓ indicates decreased risk for cognitive decline
↔ indicates no significant impact to cognitive decline
<sup>a</sup> used aggregate data that did not allow simultaneous adjustments of covariates, causality cannot be inferred
<sup>e</sup> reference group = vitamin K antagonists
<sup>b</sup> reference group = sulfonylureas
<sup>c</sup> reference group = metformin monotherapy
<sup>d</sup> reference group = tamoxifen

### Medical conditions

Out of the 80 articles reviewed, 39 (48.8%) examined the interplay between medical conditions and ADRD. Of them, 16 articles focused on cardiovascular and metabolic conditions, 8 addressed infections, inflammatory and immune-related conditions, 7 articles tackled neurological/ophthalmological conditions, 5 examined issues related to physical function and frailty, 4 delved into psychiatric conditions, and the remaining 7 articles investigated other risk factors, such as cancer,^30, 60^ kidney disease,^21, 51^ osteoarthritis,^54^ and hip fracture.^46^

#### Cardiovascular and metabolic conditions

A clear thread emerging from our analysis is the association of cardiovascular and metabolic conditions with an increased risk for ADRD. Diabetes, for instance, has been spotlighted in several studies.^20, 51, 54, 61, 62^ Hypoglycemia, a common complication of diabetes treatment, has also been identified as a risk factor.^34, 45, 57, 63^ Extensive research has been done on the relationship between blood pressure and ADRD using EHR data. Hypertension,^51, 62, 64, 65^ hypotension^49^ and blood pressure variability^66^ have all been shown to contribute to an increased risk for ADRD. Additional risk factors in this category include coronary artery disease,^51^ stroke,^51^ and hyperlipidemia.^51^ Obesity is contradictory with one study showing obesity as a risk factor for dementia,^38^ while another study suggesting it might have a protective effect.^25^ The age at which obesity is assessed seems to play a role, as midlife obesity has been more consistently linked with an increased risk of dementia later in life.^38^ However, obesity in older adults, especially those in their 70s and 80s, might be associated with a decreased risk of dementia.^67^ These findings in older adults may be driven by those who are underweight, who may in fact be frailer and have an association with increased dementia risk.

#### Infections, inflammatory and immune-related conditions

HIV,^68, 69^ E. coli,^70^ and Covid-19^14^ have been identified as risk factors for ADRD. Contrary to previous findings,^71^ several common infections–such as sepsis, pneumonia, other lower respiratory tract infections, urinary tract infections, and skin and soft tissue infections–were not associated with an increased risk of ADRD.^27^ A subtype of infection deserving mention is that involving herpes viruses. One study observed a slightly decreased risk of dementia in individuals with symptomatic Herpes Simplex Virus 1 (HSV-1) infections not treated by antivirals and a pronounced 25% decreased risk in those who were treated with antivirals.^71^ Another study found no overall effect of a diagnosis with Herpes Zoster (HZ) on the incidence of dementia. However, it did identify a minor protective association between HZ and dementia, an association observed exclusively in frail individuals and females, and only for mixed or unspecified dementia.^35^ Furthermore, a study reported that patients grouped within the inflammatory/autoimmune disease cluster exhibited an elevated ADRD risk.^30^ Inflammatory bowel disease was also found as a risk factor.^17^ Both high urate^25^ and gout^40^ were associated with a decreased risk for ADRD, suggesting a protective effect of uric acid. The antioxidant effects of uric acid are a potential mechanism for protective effects of obesity observed in some studies^22^.

#### Psychiatric conditions

The interplay between depression and ADRD remains unclear. Some view depression as a symptom, while others consider it as a precursor of ADRD. Of the articles included in our final analysis, three explored the relationship between depression and ADRD, with all identifying a depression as a risk factor for ADRD.^43, 54, 72^ Psychotic disorders have also been reported as a risk factor.^44^

#### Neurological/ophthalmological conditions

The eyes and brain also form crucial nodes in the ADRD risk network. Retinal vascular occlusion may be associated with an increased ADRD risk.^59^ Visual impairment, determined by visual acuity, has also been linked to an elevated ADRD risk^26^ although another study did not find this association.^73^ The impact of diabetic retinopathy, a complication from diabetes, remains ambiguous: one study indicated an increased risk,^74^ while another observed no effect.^75^ Both traumatic brain injury^76^ and epilepsy^77^ have been identified as risk factors.

#### Physical function and frailty

Frailty metrics also weigh in on the ADRD risk scales. Being underweight was identified as a risk factor for ADRD,^25, 49, 54^ except in one study.^38^ Avoidance of the increased risk associated with being underweight is one potential mechanism for the protective effect sometimes seen with obesity.^22^ Low physical function, as measured by grip strength and the Short Physical Performance Battery (SPPB), was also associated with an increased risk.^53^ Conversely, another study found that neither physical inactivity nor unintentional low caloric intake was associated with an increased ADRD risk.

#### Other medical conditions

Several studies have investigated a miscellany of medical conditions and their potential of ADRD linkages. Cancer emerges as a notable example. One study found that patients grouped within the cancer disease cluster had an elevated risk of ADRD.^30^ Conversely, another study determined that both malignant melanoma and non-melanoma skin cancers were associated with a decreased ADRD risk, suggesting a protective effect.^60^ Kidney disease,^21, 51^ hip fracture,^46^ and osteoarthritis^54^ were also identified as risk factors for ADRD.

### Medical interventions

In light of the risk posed by medical conditions to ADRD, researchers have examined various medical interventions to determine if they could mitigate the risk of ADRD. Of the 80 articles assessed, 23 (28.8%) analyzed the association between medical interventions and ADRD. Out of these, 10 were related to cardiovascular and metabolic interventions, 4 to immune, infection, and inflammatory interventions, 4 to psychiatric interventions, 3 to oncology, and 4 to other interventions.

#### Cardiovascular and metabolic-related interventions

Treatments targeting cardiovascular and metabolic conditions have been prominently investigated for their potential to reduce ADRD risk. Medications such as rosuvastatin,^78^ telmisartan,^52^ anticoagulants,^37^ and aspirin,^15^ which are primarily geared towards cardiovascular health, have all demonstrated efficacy in reducing the ADRD risk. In the context of diabetes management, metformin exhibited no association with incident dementia compared with a lack of initial treatment within the first 6 months post-diagnosis.^79^ However, it presented a mild protective effect compared to sulfonylureas.^18, 80^ Conversely, thiazolidinedione monotherapy, as well as metformin and thiazolidinedione combined therapy, both displayed a reduced ADRD risk compared to metformin monotherapy.^80^ Another study involving patients with atrial fibrillation and type 2 diabetes found that sodium-glucose co-transporter 2 inhibitors decreased the risk of dementia compared to patients not on this medication class.^16^ Among surgical interventions, bariatric surgery was found to be associated with an increased risk of ADRD,^81^ while carotid endarterectomy had no discernible impact.^55^

#### Immune, infection and inflammatory-related interventions

Tumor necrosis factor blocking agent,^82^ methotrexate^41^ and antiherpetic medications^48^ were all found to have protective effects against ADRD. While nonsteroidal anti-inflammatory drugs (NSAIDs)^36^ were observed to increase the risk.

#### Psychiatric-related interventions

Two studies had contradictory conclusions on the effect of the selective serotonin reuptake inhibitor (SSRI) antidepressant class on the subsequent risk of ADRD, with one study identifying it as a risk factor^83^ and the other a protective one.^78^ Trazodone, another serotonergic antidepressant, which is now more commonly prescribed for insomnia, was reported to be a risk factor.^39^

#### Oncology and other Interventions

Androgen deprivation therapy was found to be associated with an increased risk for ADRD by two studies published by the same team.^84, 85^ Aromatase inhibitor therapy and tamoxifen, both targeting hormone receptor-positive breast cancer, showed no evidence of a difference in dementia risk.^86^

### Lifestyle, socioeconomic, psychosocial and environmental factors

EHR data have been utilized to examine the impact of lifestyles, socioeconomic, psychosocial, and environmental factors on ADRD. Out of the articles assessed, 14 (17.5%) were related to this topic. Specifically, 5 articles focused on lifestyles, 5 examined socioeconomic factors, 3 delved into psychosocial factors, and 3 addressed environmental factors.

#### Lifestyle

Both smoking^54^ and alcohol use– as determined by overall consumption and alcohol-induced loss of consciousness^56^– were identified as risk factors for ADRD. Conversely, a healthy lifestyle, characterized by no current smoking, moderate alcohol consumption, regular physical activity, healthy diet, adequate sleep duration, less sedentary behavior, and frequent social contact, exhibited a protective effect against ADRD in patients with type II diabetes.^29^ However, diet alone was not found to be protective against ADRD.^22, 87^

#### Socioeconomic factors

Higher education is believed to have neuroprotective effects, and such effects were observed in two out of three studies analyzing the association between education and ADRD risk.^88, 89^ However, the third study found no significant correlation.^23^ Neighborhood disadvantage^54, 90^ and low occupational position^23^ were associated with higher risk of ADRD.

#### Psychosocial factors

Psychosocial factors such as social isolation have been identified as risk factors for ADRD.^31^ In contrast, frequent social contact appears to be a protective factor.^24^ Another metric, the “feeling of loneliness,” was not associated with either an increased or decreased risk.^31^

#### Environmental factors

EHR data was also used to analyze several environmental risk factors for ADRD. Being born in high stroke mortality states^91^ and exposure to Agent Orange among veterans^92^ were found to be associated with an increased risk of ADRD. Additionally, lithium levels in drinking water were associated with greater risk of dementia in women.^93^

## DISCUSSION

In this systematic review, we searched three extensive databases to identify articles related to the analysis of risk factors for dementia using EHR data. The final selection of 80 articles spans a wide range of risk factors, including medical conditions, interventions, lifestyle, socioeconomic status, psychosocial, and environmental factors. The majority of studied medical conditions were associated with an elevated risk of ADRD, whereas medical interventions addressing these conditions often reduced the ADRD risk. Using large and diverse EHR datasets has enriched the literature on antecedent risk factors for dementia and confirmed findings from smaller sample studies.

Longitudinal EHR data are suitable for ADRD research, given the characteristically slow and insidious onset of ADRD. The prolonged latency period between exposure to risk factors and the onset of clinical symptoms necessitates extended observation to accurately identify early signs and risk factors, and to determine the chronological order of events, thereby facilitating a possible assessment of causality.

Utilizing EHR datasets provides several benefits for ADRD research, especially regarding the exploration of diverse medical conditions and interventions to identify risk and protective factors for cognitive impairment and dementia. The extensive data available through EHRs furnish researchers with a wealth of variables, enabling simultaneous exploration of numerous potential risk and protective factors, allowing for comprehensive adjustments for confounders. This, in turn, yields more accurate and holistic insights into the complex aspects of ADRD research. Access to large and diverse EHR datasets not only increases statistical power, enhancing the ability to discern associations between potential risk or protective factors and cognitive decline,^94^ but also enables the research of rare events and identification of specific subgroups with unique risk profiles or disease trajectories.^60, 92^ Notably, these datasets encompass individuals from various ethnic, socio-economic, and geographical backgrounds, thereby facilitating the study across different populations and allowing the examination of various disease subtypes, trajectories, and clinical presentation variations given the significant clinical heterogeneity within ADRD. Compared to studies using claim data, clinical trials, and convenient sample observational studies, EHR offers rich, detailed clinical data that allows for in-depth studies into the clinical aspects and mechanisms of ADRD. EHR datasets can confirm findings from such studies and also offer unique insights into factors that might be overlooked or absent in them. As part of the current review, we have discovered that several factors have been studied exclusively using EHR data.

To facilitate the investigation of risk factors—including socioeconomic aspects, lifestyle, and environmental factors—EHR data are often linked to other types of datasets using patient identifiers, such as names, social security numbers, and zip codes. This approach aids in examining factors not present in the EHR and also allows for the inclusion of confounding factors from the EHR.

Utilizing EHR datasets for ADRD research has carved out a niche in contemporary scientific exploration, offering valuable insights derived from diverse and substantial patient data. Nonetheless, the navigation through this informational wealth unveils notable limitations that must be acknowledged.

One pivotal constraint revolves around the heterogeneity and quality of outcome measures across EHR-based studies. Despite the commonality of employing dementia as a focal outcome, the definition thereof remains inconsistently applied, varying in the inclusion or exclusion of vascular dementia, LBD, and diverging in the coding systems (ICD, SNOMED CT, READ) utilized for outcome definition. While some studies leverage cognitive tests like the Montreal Cognitive Assessment, they are often restricted by smaller sample sizes. Utilizing diagnostic codes like ICD for outcome measures scales up the sample size, albeit potentially compromising the specificity of dementia diagnoses.

Furthermore, the inherent biases prevalent in observational studies, characterized by potential confounding, selection bias, and information bias, remains a ubiquitous issue. EHR data, notwithstanding its vastness, often encounters challenges related to missing values, which may inadvertently skew results and interpretations. For instance, death is viewed as a competing risk for ADRD, yet this information might be absent from the EHR. While many studies have cross-referenced the EHR with external databases (e.g., state mortality files, social security death index records) to obtain death information, not all of these studies explicitly state whether they have done so.

Methodological concerns also materialize, particularly regarding statistical applications in observational ADRD studies. In observational studies on ADRD with long follow-up time, individuals are often at risk of experiencing competing events, such as death, that can preclude the occurrence of the event of interest (e.g., diagnosis of AD). The Cox model is a widely used method for survival analysis, but it is not suitable for handling competing risks appropriately, as it treats competing events as censored, which can lead to biased and inaccurate results when the assumption of independent censoring is violated. On the other hand, the Fine-Gray model allows for estimating the effect of covariates on the sub-distribution hazard, providing insight into the relationships between risk and protective factors and the event of interest while accounting for competing risks.

Most studies chose to simply adjust for confounders in the survival model. However, this method may not adequately control for confounding, especially when there are many confounders or when there is substantial overlap in covariate distributions between treatment groups. It can also lead to issues such as multicollinearity and overfitting. Under the scenario of high-dimensional confounding, propensity score methods such as propensity score matching (PSM) and inverse probability weighting (IPW) should be used to reduce the dimensionality and balance covariates between groups to reduce bias in the estimation of exposure or treatment effects. It is imperative to note that EHR-based studies, while offering valuable insights, do not conclusively establish causality due to the potential influence of uncontrolled confounding variables. Investigations into the linkage between depression and dementia exemplify this dilemma. Moreover, EHR-based studies are not randomized controlled clinical trials, which are the gold standard for establishing causality.

Lastly, a geographic and demographic constraint arises. Notwithstanding the extensive data embedded in EHR systems, much research is localized to specific healthcare systems or geographic locales, thereby limiting generalizability. Despite the widespread availability of longitudinal EHR datasets across various healthcare systems and regions, such as the Mayo Clinic and Mass General Brigham, research has largely been confined to particular EHRs, such as those in Kaiser Permanente. While the VHA dataset provides a national scope, its predominant representation of male individuals poses a demographic limitation. Consequently, the paucity of research leveraging expansive populations, especially on a national scale (e.g., via the ENACT network), underscores an imminent need for future endeavors to explore dementia risk utilizing diverse, comprehensive EHR datasets.

### Future directions

The analysis of the articles suggests several avenues for future investigation using EHR data. First, the majority of EHR-based studies to date have focused on populations with well-defined medical conditions, such as diabetes, hypertension, cancer, and HIV. There exists an opportunity— and a necessity—to broaden the scope of research to encompass a wider arrange of specific groups. These should include sexual and gender minorities,^95, 96^ indigenous populations, individuals who demonstrate resilience against cognitive decline, and those belonging to various psychiatric cohorts. Second, integrating diverse longitudinal EHR databases could expand the study population and validate findings across different institutions and geographic locations. For instance, UK has shown robust use of national datasets. In contrast, the US and other countries appear to underutilize national-level EHR datasets. The expanded use of such comprehensive data sources could provide a more representative sample and enhance the generalizability of research outcomes. Third, EHRs do not capture all relevant data, necessitating linkage with other datasets,^97^ such as Medicare records, genetic data, socioeconomical status,^98^ lifestyle, crime data, environmental factors (e.g., air pollution, wildfires, climate change, toxic chemicals). Fourth, there are considerable opportunities to investigate additional risk or protective factors that non-EHR studies have identified but were overlooked in the reviewed articles, including genetic markers (e.g., apolipoprotein E, presenilin 1 and 2, and amyloid precursor protein),^99^ environmental toxins (e.g., lead, pesticides),^100^ mild traumatic brain injury,^101^ endocrine factors (such as hypothyroidism), sleep disturbance (like sleep apnea or chronic sleep deprivation),^8, 102^ bilingualism,^103^ vitamin and nutritional deficiencies,^104^ and the microbiome (e.g., gut microbiome).^105^ Fifth, while the existing literature primary focuses on dementia or AD, fewer studies address the early onset of AD and the initial stages of cognitive decline, such as mild cognitive impairment and subjective cognitive decline. Sixth, almost all the reviewed articles have used data from structured fields of the EHR. Certain conditions and symptoms (e.g., hearing loss, sleep disturbances) that are not consistently captured in structured EHR data may require the examination of clinical notes to identify them, which often require AI and natural language processing. Finally, while a wide array of medical conditions adversely affects long-term cognitive outcomes, the clarity regarding whether receiving medical treatments would reduce risk remains obscure. Compared to the numerous studies exploring the interaction between medical conditions and ADRD, there is a scarcity of studies examining pharmacological treatment and surgical effects. Consequently, future research should pivot towards studying the association of medical interventions with cognitive decline across a more expansive area.

### Limitations

Several limitations warrant acknowledgment in this review. Firstly, our search was constrained to three databases, potentially overlooking relevant studies listed elsewhere. Secondly, our article search, confined to a limited set of terms, might have omitted pertinent articles that mentioned specific EHR components (e.g., clinical notes) or utilized alternative terminology, such as “primary care dataset,” instead of EHR. While our search terms primarily targeted titles and abstracts, the term EHR might have been mentioned in the methods section of a manuscript, which we might have overlooked. Thirdly, we abstained from conducting a bias assessment for the observational studies included, such as utilizing ROBINS-E. Given the employment of an EHR database, biases are often present in data collection and outcome measures. Fourthly, while this review meticulously explores EHR-based observational studies, it does not aim to provide a comprehensive overview of the field. Finally, we refrained from conducting a meta-analysis due to the varying confounders adjusted for in different studies, complicating cross-study comparisons.

## CONCLUSION

EHR data, with its rich and diverse longitudinal real-world information, provides substantial insights into the medical conditions, interventions, lifestyle, socioeconomic, and environmental factors associated with ADRD risk. Looking ahead, research should focus on diversifying study populations and integrating EHR data across geographical locations and with non-EHR datasets. There is also a need to enhance the extraction of information from unstructured text to explore a broader range of risk factors for ADRD.

## Data Availability

All data produced in the present work are contained in the manuscript

## Data Availability

No data was used for the research described in the article.

## CRediT authorship contribution statement

*Conceptualization: Wang, Yang*

*Data curation: Wang, Yang, Sha, Kuraszkiewicz, Leonik*

*Formal analysis: Wang, Yang, Sha, Leonik, Marshall*

*Investigation: All authors*

*Methodology: Wang, Yang*

*Project administration: Wang, Yang*

*Supervision: Wang, Marshall*

*Roles/Writing - original draft: Wang, Yang*

*Writing - review & editing: All authors*

*Funding acquisition: Wang*

## Declaration of Generative AI and AI-assisted technologies in the writing process

*During the preparation of this work the authors used GPT-4 in order to improve the readability and language. After using this tool/service, the authors reviewed and edited the content as needed and take full responsibility for the content of the publication*.

## REFERENCES

1. Organization WH. Global action plan on the public health response to dementia 2017–2025. 2017.

2. Jack Jr CR, Bennett DA, Blennow K, Carrillo MC, Dunn B, Haeberlein SB, et al. NIA-AA research framework: toward a biological definition of Alzheimer’s disease. Alzheimer’s & Dementia. 2018;14(4):535–62.

3. Sperling RA, Aisen PS, Beckett LA, Bennett DA, Craft S, Fagan AM, et al. Toward defining the preclinical stages of Alzheimer’s disease: Recommendations from the National Institute on Aging-Alzheimer’s Association workgroups on diagnostic guidelines for Alzheimer’s disease. Alzheimer’s & dementia. 2011;7(3):280–92.

4. Association As. 2023 Alzheimer’s disease facts and figures. Alzheimer’s & dementia. 2023;19(4).

5. Veitch DP, Weiner MW, Aisen PS, Beckett LA, DeCarli C, Green RC, et al. Using the Alzheimer’s Disease Neuroimaging Initiative to improve early detection, diagnosis, and treatment of Alzheimer’s disease. Alzheimers Dement. 2022;18(4):824–57. PMID: 34581485.

6. Dagley A, LaPoint M, Huijbers W, Hedden T, McLaren DG, Chatwal JP, et al. Harvard aging brain study: dataset and accessibility. Neuroimage. 2017;144:255–8.

7. Wolters FJ, Segufa RA, Darweesh SKL, Bos D, Ikram MA, Sabayan B, et al. Coronary heart disease, heart failure, and the risk of dementia: A systematic review and meta-analysis. Alzheimers Dement. 2018;14(11):1493–504. PMID: 29494808.

8. Shi L, Chen SJ, Ma MY, Bao YP, Han Y, Wang YM, et al. Sleep disturbances increase the risk of dementia: A systematic review and meta-analysis. Sleep Med Rev. 2018;40:4–16. PMID: 28890168.

9. Kuiper JS, Zuidersma M, Oude Voshaar RC, Zuidema SU, van den Heuvel ER, Stolk RP, et al. Social relationships and risk of dementia: A systematic review and meta-analysis of longitudinal cohort studies. Ageing Res Rev. 2015;22:39-57. PMID: 25956016.

10. Page MJ, McKenzie JE, Bossuyt PM, Boutron I, Hoffmann TC, Mulrow CD, et al. The PRISMA 2020 statement: an updated guideline for reporting systematic reviews. International journal of surgery. 2021;88:105906.

11. Knottnerus A, Tugwell P. STROBE--a checklist to Strengthen the Reporting of Observational Studies in Epidemiology. J Clin Epidemiol. 2008;61(4):323. PMID: 18313555.

12. Cox DR. Regression models and life-tables. Journal of the Royal Statistical Society: Series B (Methodological). 1972;34(2):187–202.

13. Fine JP, Gray RJ. A proportional hazards model for the subdistribution of a competing risk. Journal of the American statistical association. 1999;94(446):496–509.

14. Wang L, Davis PB, Volkow ND, Berger NA, Kaelber DC, Xu R. Association of COVID-19 with New-Onset Alzheimer’s Disease. J Alzheimers Dis. 2022;89(2):411–4. PMID: 35912749.

15. Gorenflo MP, Davis PB, Kendall EK, Olaker VR, Kaelber DC, Xu R. Association of Aspirin Use with Reduced Risk of Developing Alzheimer’s Disease in Elderly Ischemic Stroke Patients: A Retrospective Cohort Study. J Alzheimers Dis. 2023;91(2):697–704. PMID: 36502331.

16. Proietti R, Rivera-Caravaca JM, López-Gálvez R, Harrison SL, Marín F, Underhill P, et al. Cerebrovascular, Cognitive and Cardiac Benefits of SGLT2 Inhibitors Therapy in Patients with Atrial Fibrillation and Type 2 Diabetes Mellitus: Results from a Global Federated Health Network Analysis. J Clin Med. 2023;12(8). PMID: 37109151.

17. Aggarwal M, Alkhayyat M, Abou Saleh M, Sarmini MT, Singh A, Garg R, et al. Alzheimer Disease Occurs More Frequently In Patients With Inflammatory Bowel Disease: Insight From a Nationwide Study. J Clin Gastroenterol. 2023;57(5):501–7. PMID: 35470286.

18. Newby D, Linden AB, Fernandes M, Molero Y, Winchester L, Sproviero W, et al. Comparative effect of metformin versus sulfonylureas with dementia and Parkinson’s disease risk in US patients over 50 with type 2 diabetes mellitus. BMJ Open Diabetes Res Care. 2022;10(5). PMID: 36109050.

19. Lindbohm JV, Mars N, Walker KA, Singh-Manoux A, Livingston G, Brunner EJ, et al. Plasma proteins, cognitive decline, and 20-year risk of dementia in the Whitehall II and Atherosclerosis Risk in Communities studies. Alzheimers Dement. 2022;18(4):612–24. PMID: 34338426.

20. Barbiellini Amidei C, Fayosse A, Dumurgier J, Machado-Fragua MD, Tabak AG, van Sloten T, et al. Association Between Age at Diabetes Onset and Subsequent Risk of Dementia. Jama. 2021;325(16):1640–9. PMID: 33904867.

21. Singh-Manoux A, Oumarou-Ibrahim A, Machado-Fragua MD, Dumurgier J, Brunner EJ, Kivimaki M, et al. Association between kidney function and incidence of dementia: 10-year follow-up of the Whitehall II cohort study. Age Ageing. 2022;51(1). PMID: 35061870.

22. Akbaraly TN, Singh-Manoux A, Dugravot A, Brunner EJ, Kivimäki M, Sabia S. Association of Midlife Diet With Subsequent Risk for Dementia. Jama. 2019;321(10):957–68. PMID: 30860560.

23. Rusmaully J, Dugravot A, Moatti JP, Marmot MG, Elbaz A, Kivimaki M, et al. Contribution of cognitive performance and cognitive decline to associations between socioeconomic factors and dementia: A cohort study. PLoS Med. 2017;14(6):e1002334. PMID: 28650972.

24. Sommerlad A, Sabia S, Singh-Manoux A, Lewis G, Livingston G. Association of social contact with dementia and cognition: 28-year follow-up of the Whitehall II cohort study. PLoS Med. 2019;16(8):e1002862. PMID: 31374073.

25. Cao Z, Xu C, Yang H, Li S, Xu F, Zhang Y, et al. Associations of BMI and Serum Urate with Developing Dementia: A Prospective Cohort Study. J Clin Endocrinol Metab. 2020;105(12). PMID: 32918088.

26. Littlejohns TJ, Hayat S, Luben R, Brayne C, Conroy M, Foster PJ, et al. Visual Impairment and Risk of Dementia in 2 Population-Based Prospective Cohorts: UK Biobank and EPIC-Norfolk. J Gerontol A Biol Sci Med Sci. 2022;77(4):697–704. PMID: 34718565.

27. Muzambi R, Bhaskaran K, Rentsch CT, Smeeth L, Brayne C, Garfield V, et al. Are infections associated with cognitive decline and neuroimaging outcomes? A historical cohort study using data from the UK Biobank study linked to electronic health records. Transl Psychiatry. 2022;12(1):385. PMID: 36109502.

28. Geng T, Lu Q, Wan Z, Guo J, Liu L, Pan A, et al. Association of serum 25-hydroxyvitamin D concentrations with risk of dementia among individuals with type 2 diabetes: A cohort study in the UK Biobank. PLoS Med. 2022;19(1):e1003906. PMID: 35025861.

29. Wang B, Wang N, Sun Y, Tan X, Zhang J, Lu Y. Association of Combined Healthy Lifestyle Factors With Incident Dementia in Patients With Type 2 Diabetes. Neurology. 2022;99(21):e2336–e45. PMID: 36104282.

30. Khondoker M, Macgregor A, Bachmann MO, Hornberger M, Fox C, Shepstone L. Multimorbidity pattern and risk of dementia in later life: an 11-year follow-up study using a large community cohort and linked electronic health records. J Epidemiol Community Health. 2023;77(5):285–92. PMID: 36889910.

31. Elovainio M, Lahti J, Pirinen M, Pulkki-Råback L, Malmberg A, Lipsanen J, et al. Association of social isolation, loneliness and genetic risk with incidence of dementia: UK Biobank Cohort Study. BMJ Open. 2022;12(2):e053936. PMID: 35197341.

32. Herbert A, Wijlaars L, Zylbersztejn A, Cromwell D, Hardelid P. Data resource profile: hospital episode statistics admitted patient care (HES APC). International journal of epidemiology. 2017;46(4):1093-i.

33. Harley K, Jones C. Quality of Scottish morbidity record (SMR) data. Health bulletin. 1996;54(5):410–7.

34. Mehta HB, Mehta V, Goodwin JS. Association of Hypoglycemia With Subsequent Dementia in Older Patients With Type 2 Diabetes Mellitus. J Gerontol A Biol Sci Med Sci. 2017;72(8):1110–6. PMID: 27784724.

35. Warren-Gash C, Williamson E, Shiekh SI, Borjas-Howard J, Pearce N, Breuer JM, et al. No evidence that herpes zoster is associated with increased risk of dementia diagnosis. Ann Clin Transl Neurol. 2022;9(3):363–74. PMID: 35170873.

36. Dregan A, Chowienczyk P, Armstrong D. Patterns of anti-inflammatory drug use and risk of dementia: a matched case-control study. Eur J Neurol. 2015;22(11):1421–8. PMID: 26177125.

37. Cadogan SL, Powell E, Wing K, Wong AY, Smeeth L, Warren-Gash C. Anticoagulant prescribing for atrial fibrillation and risk of incident dementia. Heart. 2021;107(23):1898–904. PMID: 34645643.

38. Floud S, Simpson RF, Balkwill A, Brown A, Goodill A, Gallacher J, et al. Body mass index, diet, physical inactivity, and the incidence of dementia in 1 million UK women. Neurology. 2020;94(2):e123–e32. PMID: 31852815.

39. Brauer R, Lau WCY, Hayes JF, Man KKC, Osborn DPJ, Howard R, et al. Trazodone use and risk of dementia: A population-based cohort study. PLoS Med. 2019;16(2):e1002728. PMID: 30721226.

40. Lu N, Dubreuil M, Zhang Y, Neogi T, Rai SK, Ascherio A, et al. Gout and the risk of Alzheimer’s disease: a population-based, BMI-matched cohort study. Ann Rheum Dis. 2016;75(3):547–51. PMID: 25739830.

41. Newby D, Prieto-Alhambra D, Duarte-Salles T, Ansell D, Pedersen L, van der Lei J, et al. Methotrexate and relative risk of dementia amongst patients with rheumatoid arthritis: a multi-national multi-database case-control study. Alzheimers Res Ther. 2020;12(1):38. PMID: 32252806.

42. Ford AH, Yeap BB, Flicker L, Hankey GJ, Chubb SAP, Golledge J, et al. Sex hormones and incident dementia in older men: The health in men study. Psychoneuroendocrinology. 2018;98:139–47. PMID: 30144781.

43. Almeida OP, Hankey GJ, Yeap BB, Golledge J, Flicker L. Depression as a risk factor for cognitive impairment in later life: the Health In Men cohort study. Int J Geriatr Psychiatry. 2016;31(4):412–20. PMID: 26280254.

44. Almeida OP, Ford AH, Hankey GJ, Yeap BB, Golledge J, Flicker L. Risk of dementia associated with psychotic disorders in later life: the health in men study (HIMS). Psychol Med. 2019;49(2):232–42. PMID: 29564993.

45. Lee CH, Lui DTW, Cheung CYY, Woo YC, Fong CHY, Yuen MMA, et al. Different glycaemia-related risk factors for incident Alzheimer’s disease in men and women with type 2 diabetes-A sex-specific analysis of the Hong Kong diabetes database. Diabetes Metab Res Rev. 2021;37(6):e3401. PMID: 32870568.

46. Hsu WWQ, Zhang X, Sing CW, Li GHY, Tan KCB, Kung AWC, et al. Hip Fracture as a Predictive Marker for the Risk of Dementia: A Population-Based Cohort Study. J Am Med Dir Assoc. 2022;23(10):1720 e1-e9. PMID: 35988591.

47. Cui S, Fang F, Cui P, Jiang Q, Xu S, Xu Z, et al. Associations between the use of beta-adrenoceptor acting drugs and the risk of dementia in older population. Front Neurol. 2022;13:999666. PMID: 36619918.

48. Schnier C, Janbek J, Williams L, Wilkinson T, Laursen TM, Waldemar G, et al. Antiherpetic medication and incident dementia: Observational cohort studies in four countries. Eur J Neurol. 2021;28(6):1840–8. PMID: 33657269.

49. Perera G, Rijnbeek PR, Alexander M, Ansell D, Avillach P, Duarte-Salles T, et al. Vascular and metabolic risk factor differences prior to dementia diagnosis: a multidatabase case-control study using European electronic health records. BMJ Open. 2020;10(11):e038753. PMID: 33191253.

50. Hafdi M, Hoevenaar-Blom MP, Beishuizen CRL, Moll van Charante EP, Richard E, van Gool WA. Association of Benzodiazepine and Anticholinergic Drug Usage With Incident Dementia: A Prospective Cohort Study of Community-Dwelling Older Adults. J Am Med Dir Assoc. 2020;21(2):188–93.e3. PMID: 31300339.

51. Kuo SC, Lai SW, Hung HC, Muo CH, Hung SC, Liu LL, et al. Association between comorbidities and dementia in diabetes mellitus patients: population-based retrospective cohort study. J Diabetes Complications. 2015;29(8):1071–6. PMID: 26233574.

52. Liu CH, Sung PS, Li YR, Huang WK, Lee TW, Huang CC, et al. Telmisartan use and risk of dementia in type 2 diabetes patients with hypertension: A population-based cohort study. PLoS Med. 2021;18(7):e1003707. PMID: 34280191.

53. Papadopoulos E, Abu Helal A, Berger A, Jin R, Romanovsky L, Monginot S, et al. Objective measures of physical function and their association with cognitive impairment in older adults with cancer prior to treatment. J Geriatr Oncol. 2022;13(8):1141–8. PMID: 35879200.

54. Pham ANQ, Lindeman C, Voaklander D, Wagg A, Drummond N. Risk factors for incidence of dementia in primary care practice: a retrospective cohort study in older adults. Fam Pract. 2022;39(3):406–12. PMID: 34910126.

55. Halliday A, Sneade M, Björck M, Pendlebury ST, Bulbulia R, Parish S, et al. Editor’s Choice - Effect of Carotid Endarterectomy on 20 Year Incidence of Recorded Dementia: A Randomised Trial. Eur J Vasc Endovasc Surg. 2022;63(4):535–45. PMID: 35272949.

56. Kivimäki M, Singh-Manoux A, Batty GD, Sabia S, Sommerlad A, Floud S, et al. Association of Alcohol-Induced Loss of Consciousness and Overall Alcohol Consumption With Risk for Dementia. JAMA Netw Open. 2020;3(9):e2016084. PMID: 32902651.

57. Hendrie HC, Zheng M, Li W, Lane K, Ambuehl R, Purnell C, et al. Glucose level decline precedes dementia in elderly African Americans with diabetes. Alzheimers Dement. 2017;13(2):111–8. PMID: 27793691.

58. Lee CS, Gibbons LE, Lee AY, Yanagihara RT, Blazes MS, Lee ML, et al. Association Between Cataract Extraction and Development of Dementia. JAMA Intern Med. 2022;182(2):134–41. PMID: 34870676.

59. Lee CS, Lee ML, Gibbons LE, Yanagihara RT, Blazes M, Kam JP, et al. Associations Between Retinal Artery/Vein Occlusions and Risk of Vascular Dementia. J Alzheimers Dis. 2021;81(1):245–53. PMID: 33749651.

60. Ibler E, Tran G, Orrell KA, Serrano L, Majewski S, Sable KA, et al. Inverse association for diagnosis of Alzheimer’s disease subsequent to both melanoma and non-melanoma skin cancers in a large, urban, single-centre, Midwestern US patient population. J Eur Acad Dermatol Venereol. 2018;32(11):1893–6. PMID: 29573497.

61. Doney ASF, Bonney W, Jefferson E, Walesby KE, Bittern R, Trucco E, et al. Investigating the Relationship Between Type 2 Diabetes and Dementia Using Electronic Medical Records in the GoDARTS Bioresource. Diabetes Care. 2019;42(10):1973–80. PMID: 31391202.

62. Nagar SD, Pemu P, Qian J, Boerwinkle E, Cicek M, Clark CR, et al. Investigation of hypertension and type 2 diabetes as risk factors for dementia in the All of Us cohort. Sci Rep. 2022;12(1):19797. PMID: 36396674.

63. Hendrie HC, Zheng M, Lane KA, Ambuehl R, Purnell C, Li S, et al. Changes of glucose levels precede dementia in African-Americans with diabetes but not in Caucasians. Alzheimers Dement. 2018;14(12):1572–9. PMID: 29678640.

64. Gilsanz P, Mayeda ER, Glymour MM, Quesenberry CP, Mungas DM, DeCarli C, et al. Female sex, early-onset hypertension, and risk of dementia. Neurology. 2017;89(18):1886–93. PMID: 28978656.

65. Abell JG, Kivimäki M, Dugravot A, Tabak AG, Fayosse A, Shipley M, et al. Association between systolic blood pressure and dementia in the Whitehall II cohort study: role of age, duration, and threshold used to define hypertension. Eur Heart J. 2018;39(33):3119–25. PMID: 29901708.

66. Whiteley WN, Gupta AK, Godec T, Rostamian S, Whitehouse A, Mackay J, et al. Long-Term Incidence of Stroke and Dementia in ASCOT. Stroke. 2021;52(10):3088–96. PMID: 34192893.

67. Qizilbash N, Gregson J, Johnson ME, Pearce N, Douglas I, Wing K, et al. BMI and risk of dementia in two million people over two decades: a retrospective cohort study. Lancet Diabetes Endocrinol. 2015;3(6):431–6. PMID: 25866264.

68. Bobrow K, Xia F, Hoang T, Valcour V, Yaffe K. HIV and risk of dementia in older veterans. Aids. 2020;34(11):1673–9. PMID: 32701576.

69. Lam JO, Hou CE, Hojilla JC, Anderson AN, Gilsanz P, Alexeeff SE, et al. Comparison of dementia risk after age 50 between individuals with and without HIV infection. AIDS. 2021;35(5):821–8. PMID: 33394681.

70. Katz J, Gao H. The Alzheimer-E. coli Axis: What Can We Learn from an Electronic Health Record Platform. J Alzheimers Dis. 2021;84(2):717–21. PMID: 34569963.

71. Young-Xu Y, Powell EI, Zwain GM, Yazdi MT, Gui J, Shiner B. Symptomatic Herpes Simplex Virus Infection and Risk of Dementia in US Veterans: a Cohort Study. Neurotherapeutics. 2021;18(4):2458–67. PMID: 34244925.

72. Gilsanz P, Schnaider Beeri M, Karter AJ, Quesenberry CP, Jr., Adams AS, Whitmer RA. Depression in type 1 diabetes and risk of dementia. Aging Ment Health. 2019;23(7):880–6. PMID: 29634288.

73. Chan AX, Bakhoum CY, Bangen KJ, Bakhoum MF. Relationship between Retinal Vascular Occlusions and Cognitive Dementia in a Large Cross-Sectional Cohort. Am J Ophthalmol. 2021;226:201–5. PMID: 33529587.

74. Exalto LG, Biessels GJ, Karter AJ, Huang ES, Quesenberry CP, Jr., Whitmer RA. Severe diabetic retinal disease and dementia risk in type 2 diabetes. J Alzheimers Dis. 2014;42 Suppl 3(0 3):S109–17. PMID: 24625797.

75. Rodill LG, Exalto LG, Gilsanz P, Biessels GJ, Quesenberry CP, Jr., Whitmer RA. Diabetic Retinopathy and Dementia in Type 1 Diabetes. Alzheimer Dis Assoc Disord. 2018;32(2):125–30. PMID: 29261519.

76. Barnes DE, Kaup A, Kirby KA, Byers AL, Diaz-Arrastia R, Yaffe K. Traumatic brain injury and risk of dementia in older veterans. Neurology. 2014;83(4):312–9. PMID: 24966406.

77. Kim Y, Lhatoo S, Zhang GQ, Chen L, Jiang X. Temporal phenotyping for transitional disease progress: An application to epilepsy and Alzheimer’s disease. J Biomed Inform. 2020;107:103462. PMID: 32562896.

78. Xu J, Wang F, Zang C, Zhang H, Niotis K, Liberman AL, et al. Comparing the effects of four common drug classes on the progression of mild cognitive impairment to dementia using electronic health records. Sci Rep. 2023;13(1):8102. PMID: 37208478.

79. Salas J, Morley JE, Scherrer JF, Floyd JS, Farr SA, Zubatsky M, et al. Risk of incident dementia following metformin initiation compared with noninitiation or delay of antidiabetic medication therapy. Pharmacoepidemiol Drug Saf. 2020;29(6):623–34. PMID: 32363681.

80. Tang X, Brinton RD, Chen Z, Farland LV, Klimentidis Y, Migrino R, et al. Use of oral diabetes medications and the risk of incident dementia in US veterans aged ≥60 years with type 2 diabetes. BMJ Open Diabetes Res Care. 2022;10(5). PMID: 36220195.

81. Kim J, Kelley J, Kleinschmit K, Richards N, Adams T. Development of dementia in patients who underwent bariatric surgery. Surg Endosc. 2023;37(5):3507–21. PMID: 36581785.

82. Zhou M, Xu R, Kaelber DC, Gurney ME. Tumor Necrosis Factor (TNF) blocking agents are associated with lower risk for Alzheimer’s disease in patients with rheumatoid arthritis and psoriasis. PLoS One. 2020;15(3):e0229819. PMID: 32203525.

83. Wang C, Gao S, Hendrie HC, Kesterson J, Campbell NL, Shekhar A, et al. Antidepressant Use in the Elderly Is Associated With an Increased Risk of Dementia. Alzheimer Dis Assoc Disord. 2016;30(2):99–104. PMID: 26295747.

84. Nead KT, Gaskin G, Chester C, Swisher-McClure S, Dudley JT, Leeper NJ, et al. Androgen Deprivation Therapy and Future Alzheimer’s Disease Risk. J Clin Oncol. 2016;34(6):566–71. PMID: 26644522.

85. Nead KT, Gaskin G, Chester C, Swisher-McClure S, Leeper NJ, Shah NH. Association Between Androgen Deprivation Therapy and Risk of Dementia. JAMA Oncol. 2017;3(1):49–55. PMID: 27737437.

86. Bromley SE, Matthews A, Smeeth L, Stanway S, Bhaskaran K. Risk of dementia among postmenopausal breast cancer survivors treated with aromatase inhibitors versus tamoxifen: a cohort study using primary care data from the UK. J Cancer Surviv. 2019;13(4):632–40. PMID: 31321612.

87. Flores AC, Jensen GL, Mitchell DC, Na M, Wood GC, Still CD, et al. Prospective Study of Diet Quality and the Risk of Dementia in the Oldest Old. Nutrients. 2023;15(5). PMID: 36904280.

88. Soh Y, Whitmer RA, Mayeda ER, Glymour MM, Peterson RL, Eng CW, et al. State-Level Indicators of Childhood Educational Quality and Incident Dementia in Older Black and White Adults. JAMA Neurol. 2023;80(4):352–9. PMID: 36780143.

89. Hayes-Larson E, Ikesu R, Fong J, Mobley TM, Gee GC, Brookmeyer R, et al. Association of Education With Dementia Incidence Stratified by Ethnicity and Nativity in a Cohort of Older Asian American Individuals. JAMA Netw Open. 2023;6(3):e231661. PMID: 36877520.

90. Becerril A, Pfoh ER, Hashmi AZ, Mourany L, Gunzler DD, Berg KA, et al. Racial, ethnic and neighborhood socioeconomic differences in incidence of dementia: A regional retrospective cohort study. J Am Geriatr Soc. 2023. PMID: 36928611.

91. Gilsanz P, Mayeda ER, Glymour MM, Quesenberry CP, Whitmer RA. Association Between Birth in a High Stroke Mortality State, Race, and Risk of Dementia. JAMA Neurol. 2017;74(9):1056–62. PMID: 28759663.

92. Martinez S, Yaffe K, Li Y, Byers AL, Peltz CB, Barnes DE. Agent Orange Exposure and Dementia Diagnosis in US Veterans of the Vietnam Era. JAMA Neurol. 2021;78(4):473–7. PMID: 33492338.

93. Duthie AC, Hannah J, Batty GD, Deary IJ, Starr JM, Smith DJ, et al. Low-level lithium in drinking water and subsequent risk of dementia: Cohort study. Int J Geriatr Psychiatry. 2023;38(3):e5890. PMID: 36747488.

94. Casey JA, Schwartz BS, Stewart WF, Adler NE. Using Electronic Health Records for Population Health Research: A Review of Methods and Applications. Annu Rev Public Health. 2016;37:61–81. PMID: 26667605.

95. Hua Y, Wang L, Nguyen V, Rieu-Werden M, McDowell A, Bates DW, et al. A deep learning approach for transgender and gender diverse patient identification in electronic health records. J Biomed Inform. 2023;147:104507. PMID: 37778672.

96. Romanelli RJ, Rosenblatt AS, Marcum ZA, Flatt JD. Cognitive Impairment in Sexual and Gender Minority Groups: A Scoping Review of the Literature. LGBT Health. 2023. PMID: 37824757.

97. DeBord DG, Carreon T, Lentz TJ, Middendorf PJ, Hoover MD, Schulte PA. Use of the "Exposome" in the Practice of Epidemiology: A Primer on -Omic Technologies. Am J Epidemiol. 2016;184(4):302–14. PMID: 27519539.

98. Kind AJH, Buckingham WR. Making Neighborhood-Disadvantage Metrics Accessible - The Neighborhood Atlas. N Engl J Med. 2018;378(26):2456–8. PMID: 29949490.

99. Armstrong RA. Risk factors for Alzheimer’s disease. Folia neuropathologica. 2019;57(2):87–105.

100. Yan D, Zhang Y, Liu L, Yan H. Pesticide exposure and risk of Alzheimer’s disease: a systematic review and meta-analysis. Sci Rep. 2016;6:32222. PMID: 27581992.

101. Clark E, Faruque S, Mutebi C, Nagirimadugu NV, Kim A, Mahendran M, et al. Investigating the relationship between mild traumatic brain injury and Alzheimer’s disease and related dementias: a systematic review. J Neurol. 2022;269(9):4635–45. PMID: 35648232.

102. Guay-Gagnon M, Vat S, Forget MF, Tremblay-Gravel M, Ducharme S, Nguyen QD, et al. Sleep apnea and the risk of dementia: A systematic review and meta-analysis. J Sleep Res. 2022;31(5):e13589. PMID: 35366021.

103. Brini S, Sohrabi HR, Hebert JJ, Forrest MRL, Laine M, Hamalainen H, et al. Bilingualism Is Associated with a Delayed Onset of Dementia but Not with a Lower Risk of Developing it: a Systematic Review with Meta-Analyses. Neuropsychol Rev. 2020;30(1):1–24. PMID: 32036490.

104. Zhou J, Sun Y, Ji M, Li X, Wang Z. Association of Vitamin B Status with Risk of Dementia in Cohort Studies: A Systematic Review and Meta-Analysis. J Am Med Dir Assoc. 2022;23(11):1826 e21–e35. PMID: 35779574.

105. Coradduzza D, Sedda S, Cruciani S, De Miglio MR, Ventura C, Nivoli A, et al. Age-Related Cognitive Decline, Focus on Microbiome: A Systematic Review and Meta-Analysis. International Journal of Molecular Sciences. 2023;24(18):13680.

106. Biondo F, Jewell A, Pritchard M, Aarsland D, Steves CJ, Mueller C, et al. Brain-age is associated with progression to dementia in memory clinic patients. Neuroimage Clin. 2022;36:103175. PMID: 36087560.

107. Russ TC, Hannah J, Batty GD, Booth CC, Deary IJ, Starr JM. Childhood Cognitive Ability and Incident Dementia: The 1932 Scottish Mental Survey Cohort into their 10th Decade. Epidemiology. 2017;28(3):361–4. PMID: 28151744.

108. Schulte PJ, Warner DO, Martin DP, Deljou A, Mielke MM, Knopman DS, et al. Association Between Critical Care Admissions and Cognitive Trajectories in Older Adults. Crit Care Med. 2019;47(8):1116–24. PMID: 31107280.

